# A scoping review of online food retail: Who’s using it, what they are buying, and what tactics are used to promote healthy or unhealthy purchases

**DOI:** 10.1101/2024.02.12.24302688

**Authors:** Alexandria E. Reimold, Marissa G. Hall, Pasquale E. Rummo, Emily W. Duffy, David Gonzalez, Lindsey Smith Taillie

## Abstract

Online grocery shopping is an increasingly common part of the retail food environment. However, existing reviews have yet to synthesize the large swath of online grocery shopping research. We searched seven databases for studies reporting empirical data on online grocery retailing and shopping. Two reviewers screened search results and extracted information from 117 manuscripts containing 122 studies. Most studies were conducted in North America, Europe, and Asia. Younger, highly educated, and higher-income individuals were most likely to use online grocery and time savings, convenience, and website usability were the most common motivators for use. Common deterrents included the inability to pick perishable items and a lack of trust in the in-store shopper. Barriers included delivery and service fees, difficulty navigating online ordering, and limited availability. Individuals were more likely to purchase bulky/heavy items and less likely to purchase impulse items and perishables like fresh produce. The most common online retail promotion was price discounting. However, shoppers reported that marketing tactics seemed less noticeable online compared to brick-and-mortar stores. Online grocery has gained popularity; nevertheless, barriers may reflect inequities in access. Research is needed to further understand how shopping online influences overall food purchases and how to ensure equitable access.

## Introduction

The context in which consumers buy food matters greatly for what they purchase. For decades, a large body of research on the retail food environment (with a particular focus on grocers and large supercenters) has shown that marketing-related elements, such as product availability, prices, promotions, and placement within the store influence what consumers purchase [1–3]. Additional research shows how the *type* of food retailer matters: different consumers shop at different store types (e.g., small stores vs. grocery stores vs. big box chains) and different store types are associated with more or less healthful purchases [4]. However, the vast majority of this research has focused on the *physical* food retail environment; in other words, brick-and-mortar stores where consumers can shop in-person.

Online grocery retailers (henceforth online grocers), have created an entirely new environment for people to purchase food in. Large brick-and-mortar stores with groceries, like Wal-Mart, also offer online grocery options with both delivery and click-and-collect (i.e., the shopper purchases food online and picks up the bundled groceries on-site) services. The online grocery sector has roots in the late 1990s and early 2000s with the online launches of ASDA and Tesco in the United Kingdom, Walmart in the United States (US), and Alibaba’s in China.

However, it wasn’t until the COVID-19 pandemic that its popularity soared [5]. Though online grocers are often extensions of large brick-and-mortar chain grocers, they differ from their brick-and-mortar locations by offering the convenience of shopping from almost anywhere and freeing up the time previously spent in stores. Furthermore, in addition to “traditional” marketing elements, the online nature of online food retail also allows for unique promotional features, such as personalized banner adds or promotions [6,7], but what these consist of and how they influence the healthfulness of purchases is not well understood.

The shift to online food retail has potential implications for disparities in the nutritional quality of what people purchase and eat. The Supplemental Nutrition Assistance Program (SNAP), the largest federal food assistance program in the US, expanded the implementation of its online purchasing pilot [8], and the US Department of Agriculture proposed a rule that would allow shoppers to use Special Supplemental Nutrition Program for Women, Infant, and Children (WIC) benefits online [9]. However, prior research indicates that individuals with low incomes face barriers when grocery shopping online, namely high-cost delivery fees [10,11]. Thus, it is important to understand the extent to which populations with low incomes, who are at a disproportionate risk for diet-related diseases, are actually using online food retail, and whether and how these settings may be influencing the healthfulness of food purchases.

To our knowledge, no reviews have summarized the broad and growing body of research on the online food retail environment. Large-scale synthesis is needed to understand who shops in online stores, how food availability, accessibility, price, and promotional strategies differ in online compared to brick-and-mortar stores, and how the online food retail environment affects the nutritional quality of purchases. The objective of this scoping review is to understand the current landscape of online grocers, specific to online grocers, supermarkets, and mass merchandisers, in the real world. The current study complements other reviews on this topic that focus on specific aspects of the online food retail environment (such as marketing practices, healthfulness, and consumer uptake) [10,12–18] by broadly synthesizing a larger body of research. This review seeks to answer 1) Who uses online food retail and why do they use it? 2) What kinds of food products are sold in online food retail and how does the availability of different food products compare to brick-and-mortar stores? And 3) What pricing, promotion, placement, and other marketing tactics are the food industry using to promote their products in the online food retail landscape?

## Materials and Methods

To synthesize the existing literature on online grocers, we conducted a scoping review through June 2021. We chose to conduct a scoping review because we aim to identify gaps and understand the scope of the literature [19]. Following the PRISMA-ScR and Joannna Briggs Institute’s recommendation, we utilized the scoping review framework proposed by Peters et al. (2020) [20]. The protocol for this review is registered at Open Science Framework (<u>10.17605/OSF.IO/86D3T</u>).

### Eligibility

We established initial inclusion and exclusion criteria based on the population, concept, context categorizations [20], opting to include studies with any population, studies that focus on online food retail, and studies from any country. We defined online grocers as grocery stores on the internet that sell a wide variety of food products. We excluded studies on retailers that did not sell a wide range of food items (i.e., online retailers that specialized in energy drinks, coffee, etc.), did not have real-world data from online food retail (e.g., intervention studies in experimental labs), or focused on online ordering from restaurants, worksite cafeterias, and specialty stores rather than grocery stores. To be eligible, studies needed to be published in peer-reviewed journals, so we also excluded conference papers and unpublished dissertations and theses. Finally, we excluded studies published in languages other than English.

### Search and Screening

To develop a comprehensive search, we collaborated with a librarian on the search string and relevant databases. The search string included the keywords/strings, “online grocery shopping”, “online food shopping”, “online grocery”, “online food store”, and “online food retailer”, and the search was performed in 7 databases: PubMed, Scopus, Web of Science, Business Source Premiere, ACM Digital Library, Google Scholar, and USDA Publications. A full list of search strings adapted to the language used in each database can be found in Supplementary Table 1.

One author (AER) imported the search results into the reference management software, Zotero (Corporation for Digital Scholarship, Vienna, Virginia, US), de-duplicated multi-occurring results, then imported the possibly relevant articles into the systematic review management platform, Covidence (Veritas Health Innovation, Melbourne, Victoria, Australia). Following the Institute of Medicine’s Guidelines [21], two reviewers screened each title and abstract in Covidence and voted to include or exclude the study. Studies in which the two reviewers did not agree (i.e., one reviewer said yes but the second reviewer said no) were discussed between the two reviewers, with a third reviewer available in case of sustained disagreement. The full text of each potentially relevant study was then screened in the same way.

### Extraction

The research team identified information to extract from each study before the review began. This list was discussed and revised as the team became aware of more relevant information and themes that surfaced from the included studies. The information extracted includes: study characteristic and methodology, characteristics of online grocery users, motivations and barriers to use, the online food retail landscape, and how online grocers changed during the COVID-19 pandemic. All co-authors reviewed and confirmed the information collected and presented.

### Data Synthesis

We identified themes and gaps in the literature consistent across the included studies. These themes include the availability of online grocery and internet access, who uses online grocers, how they use them, motivations for and barriers to using them, how online grocers compare to brick-and-mortar stores regarding product availability and pricing, how and who the food industry targets in the online food retail environment, and online grocery business models. While reviewing the literature, it became clear that barriers to online grocery use naturally split into deterrents and barriers. Deterrents (i.e., not wanting to purchase fresh produce without looking at it in person) *discourage* participants from using online grocers whereas barriers (i.e., high delivery fees and low income) *prevent* participants from using online grocers. After identifying themes, we then synthesized this data and describe it thematically.

## Results

We reviewed 787 studies in total. We excluded 296 during title and abstract screening and 374 during full-text screening. This resulted in a final sample of 117 relevant published manuscripts. Three manuscripts [22–24] included multiple studies within a single publication, resulting in 122 individual studies. Figure 1 portrays the search and screening process depicted by the Preferred Reporting Items for Systematic Reviews and Meta-Analyses extension for Scoping Reviews (PRISMA-ScR) diagram. Study characteristics, methodology, results, and implications are synthesized below.

**Figure 1.**
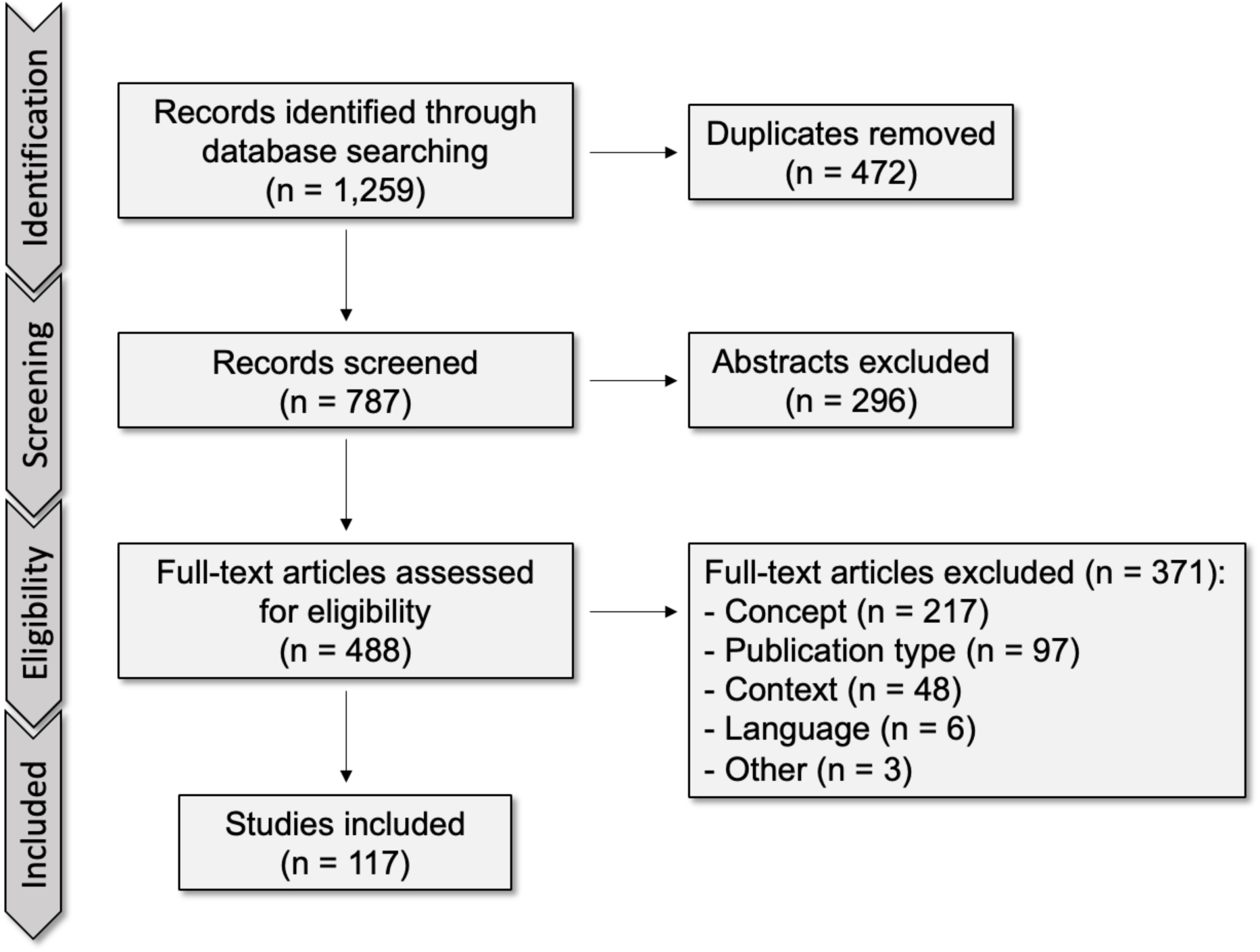
PRISMA diagram.

### Study Characteristics and Methodology

Our review represents global research over a twenty-year timespan between 2000 and 2021 (Table 1; Figure 2). Sixty-one studies were conducted in Europe, 35 in North America, 24 in Asia, three each in Africa and Oceania, one in South America, and one study [35] did not report a location. Though the oldest study included in our review was published in 2000 [25], the majority (63%) of included studies were published in the last six years of the study timespan (2016-2021). This was largely driven by the influx of 48 studies published in 2020 or later, 17 (35%) of which studied online food retail during the COVID-19 pandemic. Approximately two-thirds (64%) of included studies presented data from online food retail only (compared to both online and in-person). Authors used various methods to study online grocers including surveys (42%), consumer purchasing datasets (21%), interviews (12%), case studies (9%), store audits (8%), focus groups (4%), and content analyses (4%). Across these methods, only six (5%) studies used a mixed methods design, each of which were published in 2020 [70–75].

**Figure 2.**
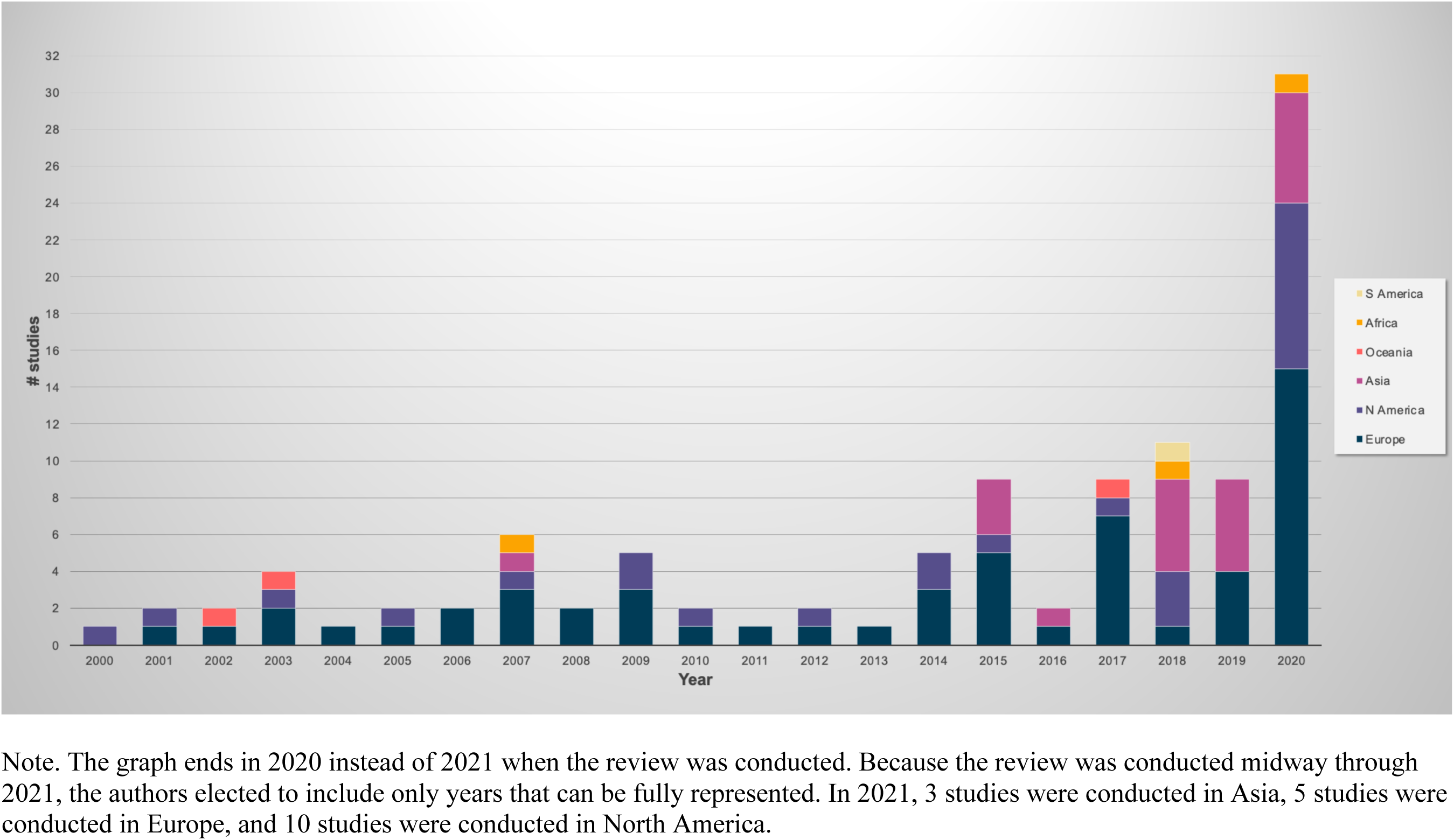
Prevalence of studies published over time by continent, 2000-2020.

**Table 1.**
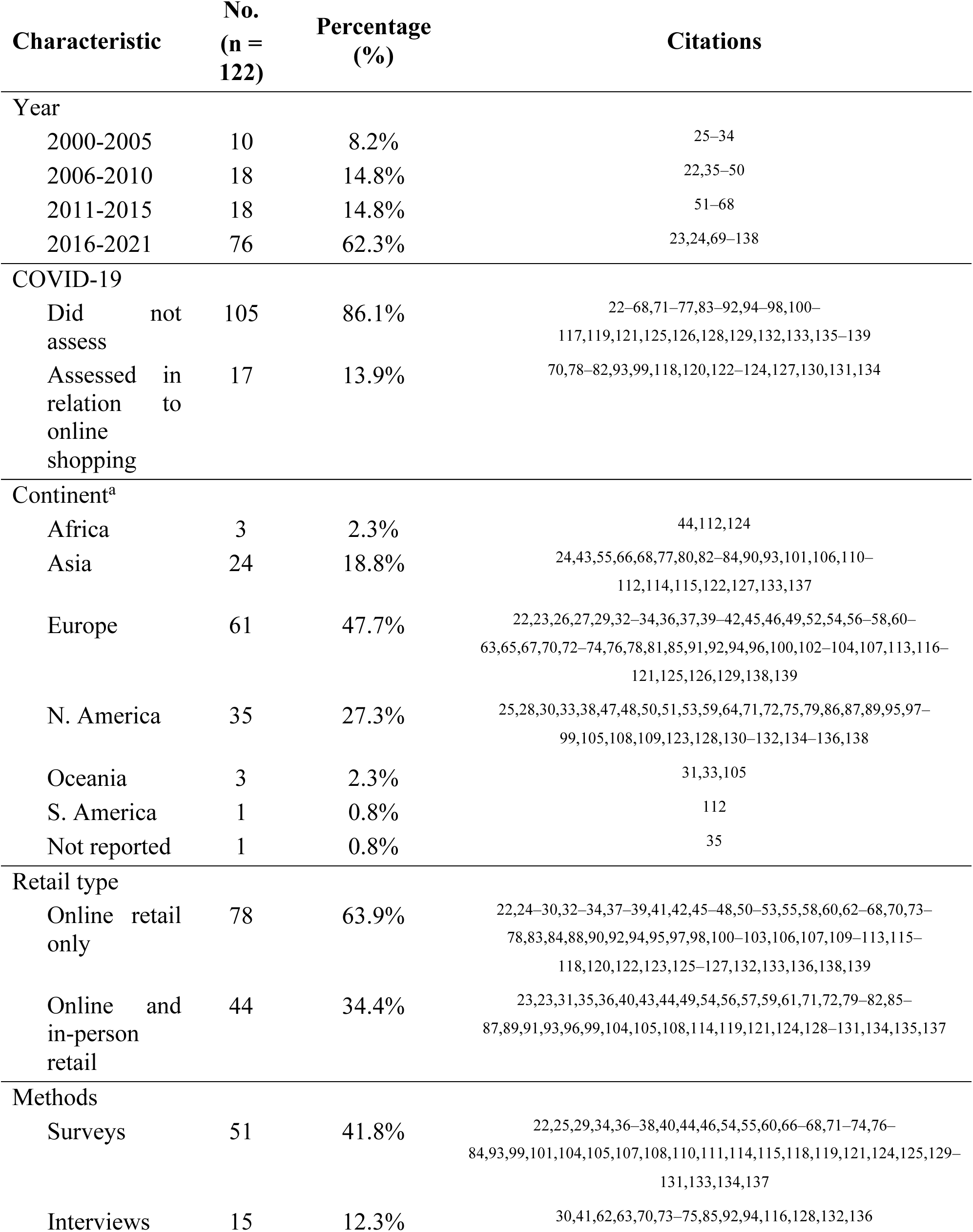
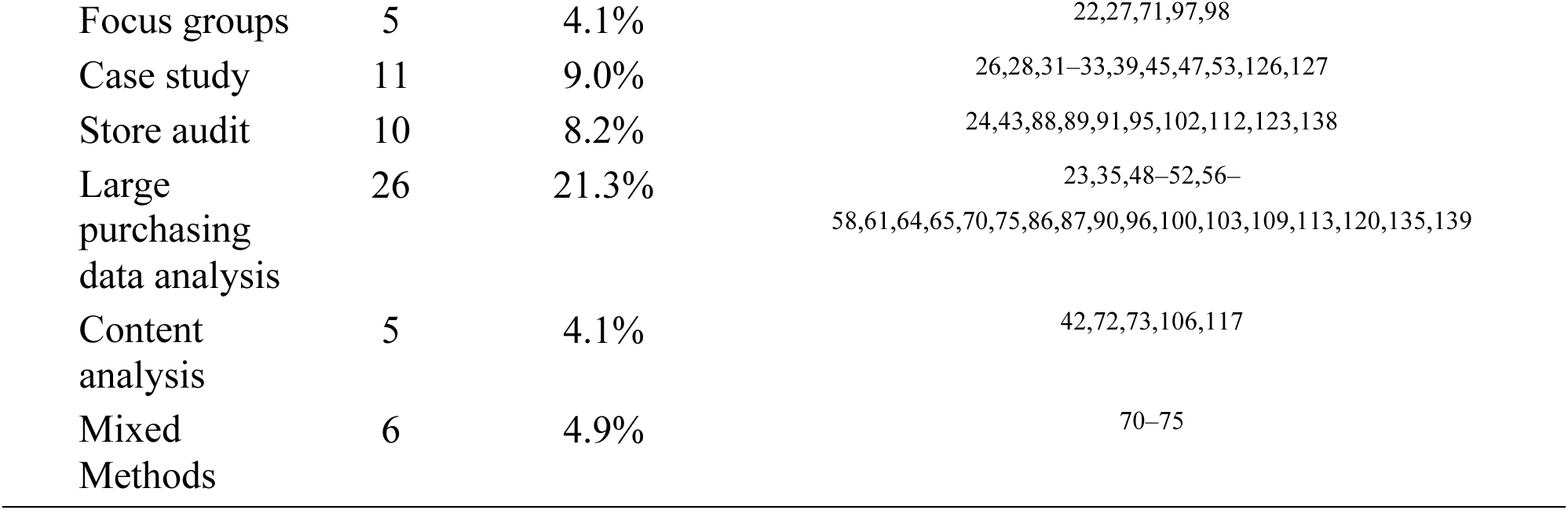
General study characteristics.

| Characteristic |  | No.<br>(n =<br>122) | Percentage<br>(%) | Citations |
| --- | --- | --- | --- | --- |
| Year |  |  |  |  |
| 2000-2005 |  | 10 | 8.2% | 25–34 |
| 2006-2010 |  | 18 | 14.8% | 22,35–50 |
| 2011-2015 |  | 18 | 14.8% | 51–68 |
| 2016-2021 |  | 76 | 62.3% | 23,24,69–138 |
| COVID-19 |  |  |  |  |
| Did not assess |  | 105 | 86.1% | 22–68,71–77,83–92,94–98,100–117,119,121,125,126,128,129,132,133,135–139 |
| Assessed in relation to online shopping |  | 17 | 13.9% | 70,78–82,93,99,118,120,122–124,127,130,131,134 |
| Continent<sup>a</sup> |  |  |  |  |
| Africa |  | 3 | 2.3% | 44,112,124 |
| Asia |  | 24 | 18.8% | 24,43,55,66,68,77,80,82–84,90,93,101,106,110–112,114,115,122,127,133,137 |
| Europe |  | 61 | 47.7% | 22,23,26,27,29,32–34,36,37,39–42,45,46,49,52,54,56–58,60–63,65,67,70,72–74,76,78,81,85,91,92,94,96,100,102–104,107,113,116–121,125,126,129,138,139 |
| N. America |  | 35 | 27.3% | 25,28,30,33,38,47,48,50,51,53,59,64,71,72,75,79,86,87,89,95,97–99,105,108,109,123,128,130–132,134–136,138 |
| Oceania |  | 3 | 2.3% | 31,33,105 |
| S. America |  | 1 | 0.8% | 112 |
| Not reported |  | 1 | 0.8% | 35 |
| Retail type |  |  |  |  |
| Online retail only |  | 78 | 63.9% | 22,24–30,32–34,37–39,41,42,45–48,50–53,55,58,60,62–68,70,73–78,83,84,88,90,92,94,95,97,98,100–103,106,107,109–113,115–118,120,122,123,125–127,132,133,136,138,139 |
| Online and in-person retail |  | 44 | 34.4% | 23,23,31,35,36,40,43,44,49,54,56,57,59,61,71,72,79–82,85–87,89,91,93,96,99,104,105,108,114,119,121,124,128–131,134,135,137 |
| Methods |  |  |  |  |
| Surveys |  | 51 | 41.8% | 22,25,29,34,36–38,40,44,46,54,55,60,66–68,71–74,76–84,93,99,101,104,105,107,108,110,111,114,115,118,119,121,124,125,129–131,133,134,137 |
| Interviews |  | 15 | 12.3% | 30,41,62,63,70,73–75,85,92,94,116,128,132,136 |

|  |  |  |  |
| --- | --- | --- | --- |
| Focus groups | 5 | 4.1% | 22,27,71,97,98 |
| Case study | 11 | 9.0% | 26,28,31–33,39,45,47,53,126,127 |
| Store audit | 10 | 8.2% | 24,43,88,89,91,95,102,112,123,138 |
| Large purchasing data analysis | 26 | 21.3% | 23,35,48–52,56–<br>58,61,64,65,70,75,86,87,90,96,100,103,109,113,120,135,139 |
| Content analysis | 5 | 4.1% | 42,72,73,106,117 |
| Mixed Methods | 6 | 4.9% | 70–75 |

### Characteristics of Online Food Retail Users

Forty-one (39%) studies reported characteristics of the individuals and households that use online grocers. These studies were published in Europe (n=18), North America (n=13), and Asia (n=10). Studies often identified younger adults as the most common group of online grocery users. However, the group “younger adults” was not defined consistently across studies, ranging anywhere between 18 and 54 years old. Though they generally followed this trend, two studies found that within young adulthood, the youngest group of adults (18-25 years old [51] and 18-30 years old [76]) used online grocers less than slightly older adults (26-45 years old [51] and 31-40 years old [76]) indicating a peak in online grocery use in middle adulthood with lower rates of use in both younger and older adulthood. Fifteen studies found that the presence of children in the household, or generally larger households, were more likely to use online grocers. However, two studies [36,52] reported that greater household size was associated with lower online grocery use. While three studies [52–54] found that being employed was associated with using online grocers, one contradictory study [77] found that unemployment was associated with using online grocers. Female gender identity, higher income, higher education, living in an urban setting, disabilities that make it difficult to leave home, limited free time, living far from a preferred store, familiarity with online shopping, frequent shopping, and large orders were all associated with greater online grocery use.

Studies from each continent identified young adults as the most common users; however, other common characteristics varied by location. Having children and/or a larger household were the most prevalent characteristics of users in European and North American studies, but this was not a widespread finding in Asian studies. Asian and European studies found that higher education was a common characteristic of online grocery users, and Asian and North American studies identified higher income as a common characteristic. North American studies were the only ones to identify disability status as a common characteristic of online grocery users.

Nineteen (15%) studies reported the frequency of online grocery use, the majority of which (n=12) were published in 2020 or 2021. Studies reported the frequency of online grocery use inconsistently and often informally (i.e., using the terms “most” or “majority” of shopping trips). From the included studies, frequency was most often reported in relation to the COVID-19 pandemic; six studies found that shopping frequency increased during this time [36,78–82]. Reportedly, online grocery shopping frequency varied as some consumers used it as often as daily while others used it as little as once per year. A 2006 European study found that the majority of online grocery users purchased food online only one to two times each year and only 10% of users purchased food online more than five times each year [36]. On the other hand, more recent studies each reported a higher prevalence of online grocery use. A 2014 North American case study of an online grocer found that, on average, customers placed three orders per month [22] and studies conducted in Asian countries in 2015, 2018, and 2020 each found that the majority of online grocery users were, at minimum, utilizing the service on a monthly basis [55,83,84]. Online grocery shopping tended to complement brick-and-mortar grocery shopping trips rather than completely replacing them [22,36,56,57,85,86]. Two European studies conducted in 2012 and 2015, in particular, found that shoppers were more likely to visit grocers in-person with online trips accounting for less than half of all grocery shopping trips among those who shop both in-person and online [56,57].

When using online grocers, individuals use various devices including computers, smartphones, and tablets. Online grocery users were more likely to use a grocery shopping list, listen to promotions or suggestions (often to buy items they had previously purchased), and plan their future meals when using online food retail as compared to in-person shopping. Studies using various methods all indicate that individuals and households tend to purchase fewer foods considered “unhealthy” or “impulse buys” when grocery shopping online [26,37,38,57,75,85,87].

### Motivations and Barriers

Studies most often reported on the motivations behind using online grocers, with almost half (49%) of the studies included in the review reporting this information (table 2). Table 3 displays specific motivations for using online grocers in detail. These motivations include time savings (n=27; 44% of studies reporting motivators) and convenience (n=24; 39% of studies reporting motivators) as the most prevalent followed by the ease of online grocery use (n=17; 28% of studies reporting motivators) and the unique variety of food products available (n=10; 16% of studies reporting motivators).

**Table 2.** Frequency of study findings reported from most to least prevalent.

| **Finding category** | **No. of studies**<br>**(n = 122)** | **Percentage (%)** |
| --- | --- | --- |
| Motivations for using online grocers | 60 | 49.2% |
| Online grocers use and socioeconomic status | 49 | 40.2% |
| Barriers and deterrents to online grocers | 45 | 36.9% |
| Characteristics of online grocery users | 41 | 33.6% |
| Proportion of shoppers who use online grocers | 41 | 33.6% |
| Online grocery pricing, promotions, and placement | 40 | 32.8% |
| Online grocery personalization | 29 | 23.8% |
| Food products sold in online grocers | 28 | 23.0% |
| Frequency of online grocery use | 24 | 19.7% |
| Online grocery availability | 22 | 18.0% |
| Online grocery business models | 11 | 9.0% |
| Online grocery user data purchasing and selling | 0 | 0% |

**Table 3.** Motivations for, deterrents, and barriers to online food retail use reported from most to least prevalent.

| **Finding** | **No. of studies**<br>**(n = 122)** | **Percentage (%)** |
| --- | --- | --- |
| Motivations for online food retail use | 60 | 49.2% |
| Time savings | 27 | 44.3% |
| Convenience | 24 | 39.3% |
| Website is easy to navigate / familiar / comfortable online | 17 | 27.9% |
| Unique food available | 10 | 16.4% |
| Good service / fast delivery / reliable | 9 | 14.8% |
| Cost efficient | 9 | 14.8% |
| Flexible hours / can shop when store is closed | 9 | 14.8% |
| Can avoid crowds / lines / parking | 9 | 14.8% |
| Good price / discounts / sales | 8 | 13.1% |
| Product quality / fresh products | 8 | 13.1% |
| Helps avoid impulse and unplanned, unhealthy purchases | 8 | 13.1% |
| Physical limitations (misc. health issues, mobility issues, cannot shop alone) | 8 | 13.1% |
| Dislike the physical store | 7 | 11.5% |
| Inclement weather (cold, rain) | 7 | 11.5% |
| Live far from preferred food retailer / no transportation / cannot drive | 7 | 11.5% |
| Lifestyle changes (childbirth, moving, getting a dog, injury, divorce) | 6 | 9.8% |
| Heavy / bulky / large number of items | 6 | 9.8% |
| No childcare | 5 | 8.2% |
| Busy schedule / provides structure | 5 | 8.2% |
| Concerns related to catching or spreading COVID-19 | 4 | 6.6% |
| Can shop around / compare prices | 4 | 6.6% |
| Gas / travel cost savings | 3 | 4.9% |
| Online grocers are available | 3 | 4.9% |
| Referral from current user / retail chain reputation | 3 | 4.9% |
| Curiosity / desire for a new experience | 1 | 1.6% |
| Accepts EBT | 1 | 1.6% |
| Deterrents to online food retail use | 40 | 32.5% |
| Unable to select own products | 23 | 57.5% |
| Lack of trust | 10 | 25% |
| Takes too much time | 6 | 15% |
| Preference for in-person shopping | 6 | 15% |
| Higher prices and lack of sales / discounts / deals | 5 | 12.5% |
| Sees in-person shopping as socialization and physical activity | 4 | 10% |
| Low perceived transaction security | 4 | 10% |
| Order and delivery problems | 4 | 10% |
| Low product assortment | 3 | 7.5% |
| Difficult to return items | 3 | 7.5% |
| Stigmatized as lazy or dependent | 1 | 2.5% |
| Requires planning / lack of spontaneity | 1 | 2.5% |
| Barriers to online food retail use | 29 | 23.6% |
| Delivery fees / minimum basket amount | 15 | 51.7% |
| Inexperience with online ordering / difficulty navigating | 9 | 31.0% |
| Online grocery / delivery availability | 4 | 13.8% |
| No smartphone or computer access | 2 | 6.9% |
| Little to no guidance available online | 2 | 6.9% |
| Online platform does not indicate WIC items | 1 | 3.4% |
| No internet or online payment access | 1 | 3.4% |

Note. Category header percentages were calculated using n=123 as the denominator. Sub-category percentages were calculated using the category header n (e.g., n=60, 40, 29) as the denominator.

In order of frequency, between 10% and 15% of studies reported that motivators included good service, particularly fast and reliable delivery; cost efficiency; ability to grocery shop even when brick-and-mortar stores are closed; opportunity to avoid crowds, long lines, and difficult parking; competitive pricing, discounts, and sales; product quality and freshness; increased ability to avoid impulse, unplanned, unhealthy purchases; physical limitations including health and mobility issues as well as an inability to travel to or navigate brick-and-mortar stores alone; distaste for shopping in-person; inclement weather such as severs cold or rain; living far from preferred food retailers, no or limited transportation, or the inability to drive one’s self. Less frequently reported motivators included recent life changes including childbirth, relocating, adopting a pet, injury, or divorce; purchasing heavy, bulky, or a large number of items; no or limited childcare; busy schedules; concerns related to catching or spreading COVID-19; desire to shop around to compare prices; gas and travel cost savings; online grocers available; referral from current users and/or the retail chain’s reputation; general curiosity and the desire for a new grocery shopping experience; ability to use EBT at checkout.

Forty (33%) studies reported on deterrents to using online grocers. Deterrents can be described as variables factored into an individuals’ choice to shop online, rather than a barrier constraining them entirely. Twenty-three (58% of studies reporting deterrents) studies described the inability of individuals to select their own food items as the most common deterrent to online grocery use. This was largely in relation to perishable items (e.g., fresh produce, meat) and product substitutions (e.g., lemons substituted for oranges). One study specifically shared that while only 35% of their survey respondents had never purchased food online, the majority of respondents (77%) had never purchased fresh produce online [84]. A second study also found that some individuals purchase their groceries online but opt to pick them up so they can select perishables in-store while saving time picking out their shelf-stable items [85]. Participants in ten (25% of studies reporting deterrents) studies shared that they did not trust stores to fulfill their orders correctly or to meet their preferences. When shopping for perishable items, individuals reported a desire to use their senses to select which items to purchase. By viewing, touching, and smelling, shoppers shared that they were more equipped to select high quality perishables that fit their preferences than store employees and were thus less inclined to purchase them online [57].

In addition to not being able to physically interact with food, other common deterrents (reported in between 10% and 15% of studies reporting deterrents) included the extra amount of time it takes to navigate online grocery platforms; general preferences for shopping in-person; higher prices and a perceived lack of discounts or deals online; viewing in-person shopping as socialization and physical activity; low perceived transaction security; and previous order and delivery issues. Additional deterrents (reported in less than 10% of studies reporting deterrents) included low product assortment; difficulty returning items; not wanting to be perceived as dependent, socially isolated, or lazy; and the planning that is often required to shop online rather than the spontaneity of stopping by a grocery store.

Twenty-nine (24%) studies reported barriers to using online grocers. Barriers, or factors outside of the shoppers’ control that keep them from using online grocers, most often included financial barriers related to delivery fees and minimum basket size requirements (n=15; 52% of studies reporting barriers), inexperience with online ordering or difficulties navigating the online food environment (n=9; 31% of studies reporting barriers) and limited availability of either online grocery or delivery (compared to click-and-collect models) (n=4; 14% of studies reporting barriers). Select studies (<10% of studies reporting barriers) additionally shared that limited smartphone or computer access, limited guidance on how to navigate online grocers, confusion around which food items are WIC approved, and limited internet access or ability to make payments online were also barriers to online grocery use.

### The Online Food Retail Landscape

Forty (33%) studies reported information on pricing, promotion, and placement tactics used in online retail or perceived by online grocery shoppers. Online grocers exhibited more price variability such that product prices fluctuated often over time and by zip code, customer demand, inventory, and both competitor and supplier pricing [88]. However, product price fluctuations in the online channel were often small [89] and overall, prices were similar to those in brick-and-mortar stores. Shoppers in one study even reported that their total basket cost when shopping online was less expensive, even when accounting for the delivery and convenience fees, largely because of the ability to easily shop around, compare prices, and find the best deals [27]. Select online grocers offer free delivery for orders above certain price thresholds [28] but this may be difficult to attain when shopping for small households or for specific items in between larger shopping trips. Other online grocers alternatively charge a monthly subscription fee that enables individuals to shop at high frequencies [28,90].

Online grocery is unique in its ability to directly attract shoppers into the store. One study conducted in Europe found that the majority of shoppers for a top online grocer found the retailer through promotions on search engines, social media, and email [120]. However, once in the online grocery platform, studies suggested that promotions may be less common compared to in brick-and-mortar stores. For example, one European study found that there were less price promotions and front-of-pack nutrition labels in the online channel of a grocer [91]. Similarly, a North American survey among online grocery shoppers found that participants wished that there were the same store circulars and in-store sales in online grocery platforms [71]. Though limited, promotional tactics included price promotions such as discounting [92] and buy-one-get-one [39]. These promotions changed weekly (as is typical in North America) or up to every eight weeks (as is more typical in Europe) [56]. Overall, online grocers are able to quickly adapt to competitively price and promote their products and retain customers in the online retail landscape.

### COVID-19 and Online Food Retail Use

Of all the studies included in this review, 17 (14%) specifically analyzed online grocery during, and/or in relation to, the COVID-19 pandemic. The COVID-19 pandemic influenced who purchased groceries online, how frequently they did it, and what they purchased. One European study found that the increase in online grocery use between January and June 2020 was larger than the increase in use in the 3 years between 2017 and 2020 [78], one North American study found that the proportion of survey participants who used online grocers almost doubled [79], and one Asian study found that one in three study participants increased their online grocery shopping during the COVID-19 pandemic [82]. Each of these studies indicate a pattern of growth in online grocery use during and/or due to the COVID-19 pandemic. An increase in online grocery use was reported across age groups; however, older individuals seemed to increase their use at a greater rate compared to younger individuals, potentially due to lower use historically. Though many studies reported a large increase in online food retail use and availability during the COVID-19 pandemic, one study in Russia found that a minority of households with high incomes were the only ones who could afford to order groceries online due to the high cost of the service [93]. Studies conducted in North America (n=6), Europe (n=5), Asia (n=5), and Africa (n=1) reported that online food retail was used during the COVID-19 pandemic due to fear of being exposed to the virus, feeling unsafe, reduced in-person retail hours, lack of public transportation, and panic buying [70,79]. Due to the pandemic, select online grocers expanded service areas; however, this growth did not significantly increase access in rural areas [70]. Along with this increase in service area, wait times also increased during the pandemic. Some online grocers implemented waitlists for new customers while others delayed food delivery by roughly 1-2 weeks [70].

## Discussion

In this scoping review of 122 relevant studies, we found that online grocery has been formally studied for over twenty years, with rapid increases at the beginning of the COVID-19 pandemic. This increase occurred as rates of online food retail availability, marketing, and use increased. Broadly, our work found that online grocery is most often used by individuals with time constraints (e.g., working parents) while other individuals face barriers to online grocery, such as delivery fees. We also found that online grocery shopping can be beneficial in some ways (i.e., fewer impulse purchases), but detrimental in others (i.e., fewer fruit and vegetable purchases) to the nutritional quality of food purchased, but that more research is needed to understand the mechanisms of how online retail influences overall food choices. Unique to this review, compared to others on the topic [10,12–18], we include studies from a wider timeframe and geography, highlighting variation in online grocery use across vast contexts.

### Study Characteristics and Methodology

Studies included in the review covered varying time periods, geographies, and methodologies. With regard to geography, most studies researched the online food environment in European countries, followed by North American, then Asian countries. This variation may reflect the different online food retail environments in each region. Online food retail first became available in Europe in 1997 with both UK-based ASDA and Tesco launching their online grocery platforms. Since then, numerous European-based grocers have launched popular online platforms and, as of 2021, six of the top 25 online retailers in Europe were online grocers [140,141]. Though to a lesser extent, compared to Europe, researchers have published on online grocery retail in North America consistently since 2000. The smaller number of publications may be because online retailers were not widely accessible in the region until 2007, with the launch of both Walmart.com and Amazon Fresh as online retailers. Asian countries first published on this topic in 2007 but did not publish frequently until 2015, likely because major online grocery platforms such as JD.com and Alibaba’s in China and BigBasket in India, along with extensions of the US based Amazon Fresh in India, Japan, and Singapore, were not available before 2011. Limited studies in this review were conducted in African, Oceanic, or South American countries. With the availability of various online grocers and delivery services in each of these geographies, this review highlights the lack of research in these areas as a major gap to be filled.

Regarding study methodology, the majority of studies used surveys to capture consumer experiences, perceptions, and behaviors followed by interviews with shoppers and analysis of large purchasing datasets. Though 26 of the 122 included studies collected some form of qualitative data, there is a distinct lack of ethnographic research and in-depth qualitative analysis. These methods could tell us more about how online grocery retail assimilates into or possibly alters existing food purchasing behaviors, how online marketing influences choices, and how efforts to increase online grocery retail accessibility are perceived and influence purchasing patterns. This review highlights the need for a more in-depth understanding of the decision-making framework related to using online grocery platforms. In addition, this review points out the inconsistent measurement of online grocery use, nutritional quality of online purchases, and the share of total household purchases that are made through online grocers. This is likely driven by researchers that primarily use surveys and interviews with self-reported measures and/or large purchasing datasets that chiefly capture data from individual retail chains. Furthermore, there is no published research leveraging multi-year longitudinal data and models that could identify changing trends in how shoppers use online grocery over time outside of the COVID-19 pandemic.

### Characteristics of Online Food Retail Users

This review revealed that young professionals, working moms, individuals with higher incomes and educational attainment, and individuals who generally want to save time are more likely to use online grocers. Each of these groups have limited time but also the financial resources or technological skills required to shop online. This generalization is supported by previous findings indicating there is a positive relationship between an individual’s perceived time pressure and their ability and willingness to pay for the convenience of online grocery shopping [142]. However, perceived time pressure is not the only reason shoppers may choose online grocers. For instance, individuals who want to avoid crowds, have disabilities that make visiting brick-and-mortar stores difficult, or want to protect themselves from getting sick particularly during the COVID-19 pandemic, are motivated to grocery shop online due to their health needs [22,25,94,95]. This motivation, combined with the increased availability of online food retail, may largely explain why the frequency of use increased over time.

The COVID-19 pandemic, as well as policy changes like the SNAP online purchasing pilot in the US, expanded online grocery use beyond individuals with higher incomes, removing some of the financial barriers to use [71]. This review, as well as other recent reviews, have highlighted barriers to equitable access to healthy foods online including high delivery fees, minimum order requirements, lack of availability of delivery services in rural areas, and inability to use WIC benefits online [10,12]. Others have highlighted concerns related to marketplace discrimination and targeted marketing of unhealthy foods in online grocers to communities with low incomes and communities of color [143]. Ensuring equitable access to healthful food in the online retail space will require a variety of community, government, and industry-led solutions, such as community delivery hubs, expansion of government approved online grocers, rigorous privacy policies for online grocers that accept federal nutrition assistance dollars, and support for companies to conduct impact assessments of their unhealthy food and beverage marketing practices [10,143,144].

### Motivations and Barriers

In this review, we found that convenience, time savings, flexibility, preference, physical access, food availability and affordability, the diverse assortment of food options, and the COVID-19 pandemic were all motivating factors for people to use online grocers. These motivators are similar to the ones found in a previous review [11] and highlight how beneficial online food retail can be for those with limited time, flexibility, physical access to grocery stores, and in times of public health crisis.

While online grocery use has become more common in recent years, this review highlighted that there are still many deterrents and barriers that need to be addressed. Individuals with limited resources may not be able to afford the cost of delivery or convenience fees [71,97] and feel a lack of control even when these fees are waived [98]. Furthermore, in rural areas, many shoppers do not have online grocery delivery services available to them and internet access is known to be unreliable [99,100,145]. Finally, many older adults prefer to shop in brick-and-mortar stores to socialize with others [146,147] and may lack the technical skills necessary to navigate online food retail websites and apps if poor health, difficulty traveling, or other situational factors lead them to need groceries delivered [146,147].

### The Online Food Retail Landscape

Studies in this review suggested that online food retail may be associated with fewer impulse food purchases. This could be due to increased meal planning when shopping online [85], the customers’ inability to interact with sensory products online [57], the delay in time between purchasing and receiving items [48,148,149], and/or marketing tactics being reportedly less salient online [57], and could have implications for both human and planetary health. Specific to human health, impulse purchases are typically comprised of hedonic foods which often contain excessive nutrients of concern such as added sugar, sodium, and saturated fat, each of which are associated with chronic disease [150–153]. Thus, the potential reduction of impulse purchases through the use of online grocers may increase the overall healthfulness of the grocery basket, possibly improving diet and health status. Specific to planetary health, list making, meal planning, and reducing impulse buys are all associated with reduced food waste [154]. The US Environmental Protection Agency cites preventing food waste as one of the most powerful steps towards limiting a households climate change footprint [155]. Therefore, online grocery shopping may be a helpful tool in the fight to improve planetary health. Future research should continue to examine the prevalence of impulse purchases in online food retail and the implications for both human and planetary health.

Though shoppers are less likely to purchase impulse items when using online food retail channels, this review, along with other recent reviews [10,11], have highlighted that people are less also likely to purchase perishable items, including fresh produce and meat, online which may have important public health implications. Specific to the US, the majority of individuals consume too few fruits, vegetables, and associated fiber, and too many ultra-processed foods high in sugar, sodium, and saturated fat [156–158]. These dietary risks are a leading cause of US deaths, decreased quality of life [159], and increased healthcare costs [160]. Further research is needed to understand the trade-offs of purchasing fewer impulse foods when shopping online at the cost of also purchasing fewer perishable foods and how this impacts overall food purchasing patterns across retailers and retail formats. For example, online grocery shopping is most often used to complement, rather than replace, brick-and-mortar grocery shopping [161,162], but limited research has studied how using online grocery platforms may change the characteristics of a shopper’s total basket, from all retail sources. One study in this review analyzed spending changes once a household started shopping online [96], but none of the 122 studies compared the healthfulness of all purchases for households that shop online and in-person to the healthfulness of all purchases for households that grocery shop exclusively in-person. Studying how the healthfulness of all food purchases change once a household begins to incorporate online grocery shopping to their food purchasing habits could identify whether the shifts in purchasing patterns related to online grocery use also shift overall purchasing, possibly leading to fewer total impulse buys and perishable food purchases across retailers and retail formats. To fully understand the potential health implications of these shopping practices, future research needs to utilize datasets that capture food purchases, both in-person and online, from a wide range of food retailers.

Findings from this review suggest that online grocers tend to manipulate prices and promotions in response to shopper and market characteristics, making them as competitive and attractive as possible. This constant manipulation and the built in best-price-possible may be why shoppers perceive online grocers as having less promotions compared to brick-and-mortar stores. As previously stated, current online grocery retail marketing is reportedly less noticeable than in-store marketing. As online grocers maintain their growth, continue to spread internationally, and discover more creative ways to market products, it will be necessary to keep monitoring their promotional activities to prevent aggressive food industry marketing of ultra-processed foods to consumers [163].

Existing reviews of marketing practices in online grocers suggests that nudging interventions related to marketing and promotion, such as suggested food swaps for products with healthier characteristics than the ones selected by the shopper, food labeling, and default options, may improve the healthfulness of online food purchases [12,17]. In a recent randomized trial, the default option of a prefilled shopping cart with fruit and vegetable items led to a significant increase in fruit and vegetable purchases made online [164]. However, further research is needed to test various nudging interventions in realistic online settings.

Limitations of this review include the exclusion of grey literature and heterogeneity of included studies. There may be relevant studies that were not peer-reviewed and thus not included in this review; however, by including only peer-reviewed studies, this search and review is highly replicable and likely includes higher quality studies. Because the purpose of this review was to assess the entire scope of literature on online grocers, the included studies vary widely in scope and research question, preventing us from meta-analyzing results.

## Conclusions

Our review indicated that research on online grocers is rapidly increasing concurrently with a global increase in online grocery availability, although deterrents and barriers to use persist. Existing studies found that individuals with higher education and income were most likely to use online food retail, largely for the convenience and time savings it provided. The most common deterrent was the inability to pick out perishable items like fresh produce and the most common barriers included delivery and service fees and difficulty navigating online ordering. This review revealed major gaps in the existing research including: research in African, Oceanic, or South American countries; more in-depth qualitative research on decision-making in real-world online food retail platforms; the prevalence of impulse food purchases online and their implications for human and planetary health; how purchasing food online alters all food purchases; and evaluation of real-world efforts to reduce the barriers to use.

Policy- and decision-makers can play a role in improving online retail by increasing internet accessibility, incentivizing retailers to reduce delivery fees, and introducing food delivery options to rural areas. Future studies could examine whether these changes to reduce barriers would increase use of online grocers among these groups of public health importance.

## Data Availability

All data produced in the present work are contained in the manuscript

---

**Supplementary Table 1.** Search strings adapted for each database searched.

| Database | Search String |
| --- | --- |
| Pubmed | “online grocery shopping” OR “online food shopping” OR “online grocery” OR “online food store” OR “online food retail” |
| Scopus | TITLE-ABS-KEY (“online grocery shopping” OR “online food shopping” OR “online grocery” OR “online food store” OR “online food retail”) |
| Web of Science | TS=(“online grocery shopping” OR “online food shopping” OR “online grocery” OR “online food retail”) |
| Business Source Premier | AB “online grocery shopping” OR AB “online food shopping” OR AB “online grocery” OR AB “online food store” OR AB “online food retail” |
| ACM Digital Library | [All: “online grocery shopping”] OR [All: "online food shopping"] OR [All: "online grocery"] OR [All: "online food store"] OR [All: "online food retail"] |
| Google Scholar | “online grocery shopping” OR “online food shopping” OR “online grocery” OR “online food store” OR “online food retail” |
| USDA Publications | “online grocery shopping” OR “online food shopping” OR “online grocery” OR “online food store” OR “online food retail” |

**Note.** The Google Scholar search included only the first 100 results as advised by a research librarian.

## Author Contributions

Conceptualization, M.G.H., P.E.R. and L.S.T.; methodology, A.E.R. and L.S.T.; validation, L.S.T.; formal analysis, A.E.R., D.G., and L.S.T.; investigation, A.E.R., E.W.D., and D.G.; data curation, A.E.R., E.W.D., D.G.; writing—original draft preparation, A.E.R. and L.S.T.; writing—review and editing, A.E.R., M.G.H., P.E.R., E.W.D., D.G., and L.S.T.; visualization, A.E.R.; supervision, L.S.T.; project administration, A.E.R. and L.S.T.; funding acquisition, M.G.H., P.E.R., L.S.T. All authors have read and agreed to the current version of this manuscript.

## Funding

This research was funded by Healthy Eating Research under Award Number 2834133. The content is solely the responsibility of the authors and does not necessarily represent the official views of healthy Eating Research.

## Acknowledgments

We thank Emily Jones for consultation while developing our search strategy and throughout this research. We also thank Catie Drawdy for assistance during the screening and extraction process.

## Conflicts of Interest

The authors declare no conflict of interest.

## Notes

**Conflicts of Interest:** We have no conflicts of interest to disclose.

**Funding:** Research reported in this manuscript was supported by Healthy Eating Research under Award Number 2834133. The content is solely the responsibility of the authors and does not necessarily represent the official views of healthy Eating Research.

### Competing Interest Statement

The authors have declared no competing interest.

## References

1. Cameron AJ, Charlton E, Ngan WW, Sacks G. A Systematic Review of the Effectiveness of Supermarket-Based Interventions Involving Product, Promotion, or Place on the Healthiness of Consumer Purchases. Curr Nutr Rep. 2016;5(3):129–138. doi:10.1007/s13668-016-0172-8

2. Castro IA, Majmundar A, Williams CB, Baquero B. Customer Purchase Intentions and Choice in Food Retail Environments: A Scoping Review. Int J Environ Res Public Health. 2018;15(11):2493. doi:10.3390/ijerph15112493

3. Caspi CE, Sorensen G, Subramanian SV, Kawachi I. The local food environment and diet: A systematic review. Health Place. 2012;18(5):1172–1187. doi:10.1016/j.healthplace.2012.05.006

4. Stern D, Ng SW, Popkin BM. The nutrient content of US household food purchases by store types. Am J Prev Med. 2016;50(2):180–190. doi:10.1016/j.amepre.2015.07.025

5. Predicting the online shopper shifts driving e-commerce momentum. NIQ. Published December 10, 2020. Accessed November 27, 2023. https://nielseniq.com/global/en/insights/education/2020/predicting-the-online-shopper-shifts-driving-e-commerce-momentum/

6. Khandpur N, Zatz LY, Bleich SN, et al. Supermarkets in Cyberspace: A Conceptual Framework to Capture the Influence of Online Food Retail Environments on Consumer Behavior. Int J Environ Res Public Health. 2020;17(22):8639. doi:10.3390/ijerph17228639

7. Yuan M, Pavlidis Y, Jain M, Caster K. Walmart Online Grocery Personalization: Behavioral Insights and Basket Recommendations. Vol 9975 LNCS.; 2016:64. doi:10.1007/978-3-319-47717-6_5

8. Stores Accepting SNAP Online | Food and Nutrition Service. Accessed August 3, 2022. https://www.fns.usda.gov/snap/online-purchasing-pilot

9. WIC Online Ordering and Transactions Proposed Rule Q&As | Food and Nutrition Service. Accessed November 27, 2023. https://www.fns.usda.gov/wic/online-ordering-transaction-proposed-rule-qas

10. Trude ACB, Lowery CM, Ali SH, Vedovato GM. An equity-oriented systematic review of online grocery shopping among low-income populations: implications for policy and research. Nutr Rev. Published online January 24, 2022:nuab122. doi:10.1093/nutrit/nuab122

11. Jilcott Pitts SB, Ng SW, Blitstein JL, Gustafson A, Niculescu M. Online grocery shopping: promise and pitfalls for healthier food and beverage purchases. Public Health Nutr. 2018;21(18):3360–3376. doi:10.1017/s1368980018002409

12. Hodges L, Lowery CM, Patel P, McInnis J, Zhang Q. A Systematic Review of Marketing Practices Used in Online Grocery Shopping: Implications for WIC Online Ordering. Nutrients. 2023;15(2):446. doi:10.3390/nu15020446

13. Le HTK, Carrel AL, Shah H. Impacts of online shopping on travel demand: a systematic review. Transp Rev. 2022;42(3):273–295. doi:10.1080/01441647.2021.1961917

14. Maganja D, Miller M, Trieu K, Scapin T, Cameron A, Wu JHY. Evidence Gaps in Assessments of the Healthiness of Online Supermarkets Highlight the Need for New Monitoring Tools: a Systematic Review. Curr Atheroscler Rep. Published online February 9, 2022. doi:10.1007/s11883-022-01004-y

15. Prabowo H, Hindarwati EN, Yuniarty. Online grocery shopping adoption: A systematic literature review. In: ; 2020:40–45. doi:10.1109/ICIMTech50083.2020.9211241

16. Tyrväinen O, Karjaluoto H. Online grocery shopping before and during the COVID-19 pandemic: A meta-analytical review. Telemat Inform. 2022;71:101839. doi:10.1016/j.tele.2022.101839

17. Valenčič E, Beckett E, Collins CE, Koroušić Seljak B, Bucher T. Digital nudging in online grocery stores: A scoping review on current practices and gaps. Trends Food Sci Technol. 2023;131:151–163. doi:10.1016/j.tifs.2022.10.018

18. Shroff A, Kumar S, Martinez LM, Pandey N. From clicks to consequences: a multi-method review of online grocery shopping. Electron Commer Res. Published online October 23, 2023. doi:10.1007/s10660-023-09761-x

19. Munn Z, Peters MDJ, Stern C, Tufanaru C, McArthur A, Aromataris E. Systematic review or scoping review? Guidance for authors when choosing between a systematic or scoping review approach. BMC Med Res Methodol. 2018;18(1):143. doi:10.1186/s12874-018-0611-x

20. M, Godfrey C, McInerney P, Munn Z, Trico A, Khalil H. Chapter 11: Scoping Reviews. In: Aromataris E, Munn Z, eds. JBI Manual for Evidence Synthesis. JBI; 2020. doi:10.46658/JBIMES-20-12

21. Institute of Medicine (US) Committee on Standards for Systematic Reviews of Comparative Effectiveness Research. Finding What Works in Health Care: Standards for Systematic Reviews. (Eden J, Levit L, Berg A, Morton S, eds.). National Academies Press (US); 2011. Accessed March 23, 2021. http://www.ncbi.nlm.nih.gov/books/NBK209518/

22. Hand C, Riley FD, Harris P, Singh J, Rettie R. Online grocery shopping: The influence of situational factors. Eur J Mark. 2009;43(9):1205–1219. doi:10.1108/03090560910976447

23. Huyghe E, Verstraeten J, Geuens M, Van Kerckhove A. Clicks as a healthy alternative to bricks: How online grocery shopping reduces vice purchases. J Mark Res. 2017;54(1):61–74. doi:10.1509/jmr.14.0490

24. Zou P, Liu J. How nutrition information influences online food sales. J Acad Mark Sci. 2019;47(6):1132–1150. doi:10.1007/s11747-019-00668-4

25. Morganosky MA, Cude BJ. Consumer response to online grocery shopping. Int J Retail Distrib Manag. 2000;28(1):17–26. doi:10.1108/09590550010306737

26. Anckar B, Walden P, Jelassi T. Creating customer value in online grocery shopping. Int J Retail Distrib Manag. 2002;30(4):211–220. doi:10.1108/09590550210423681

27. Ramus K, Nielsen NA. Online grocery retailing: What do consumers think? Internet Res. 2005;15(3):335–352. doi:10.1108/10662240510602726

28. Sheng ML. The wired mother. Technovation. 2005;25(9):1071-1077. doi:10.1016/j.technovation.2004.02.009

29. Bevan J, Murphy R. The nature of value created by UK online grocery retailers. Int J Consum Stud. 2001;25(4):279–289. doi:10.1046/j.1470-6431.2001.00195.x

30. Corbett JJ. Is online grocery shopping increasing in strength? J Food Distrib Res. 2001;32(856-2016-57733):37–40.

31. Murphy A. The emergence of online food retailing: A stakeholder perspective. Tijdschr Voor Econ En Soc Geogr. 2002;93(1):47–61. doi:10.1111/1467-9663.00182

32. Delaney-Klinger K, Boyer KK, Frohlich M. The return of online grocery shopping: A comparative analysis of Webvan and Tesco’s operational methods. TQM Mag. 2003;15(3):187–196. doi:10.1108/09544780310469334

33. Murphy AJ. (Re)solving space and time: Fulfilment issues in online grocery retailing. Environ Plan A. 2003;35(7):1173–1200. doi:10.1068/a35102

34. Chandon P, Morwitz VG, Reinartz WJ. The short-And long-term effects of measuring intent to repurchase. J Consum Res. 2004;31(3):566–572. doi:10.1086/425091

35. Khan R, Lewis M, Singh V. Dynamic customer management and the value of one-to-one marketing. Mark Sci. 2009;28(6):1063–1079. doi:10.1287/mksc.1090.0497

36. Adamides G, Marianthi G, Savvides S. Traditional Vs online attitudes towards grocery shopping in Cyprus. In: ; 2006:60-65. https://www.scopus.com/inward/record.uri?eid=2-s2.0-58249112250&partnerID=40&md5=a2aadb5c506920634b039346bd4d5499

37. Clark L, Wright P. Off Their Trolley — Understanding Online Grocery Shopping Behaviour. In: Venkatesh A, Gonsalves T, Monk A, Buckner K, eds. Home Informatics and Telematics: ICT for The Next Billion. IFIP — The International Federation for Information Processing. Springer US; 2007:157–170. doi:10.1007/978-0-387-73697-6_12

38. Gorin AA, Raynor HA, Niemeier HM, Wing RR. Home grocery delivery improves the household food environments of behavioral weight loss participants: results of an 8-week pilot study. Int J Behav Nutr Phys Act. 2007;4:58. doi:10.1186/1479-5868-4-58

39. Enders A, Jelassi T. LEVERAGING MULTICHANNEL RETAILING: THE EXPERIENCE OF TESCO.COM. Mis Q Exec. 2009;8(2):89–100.

40. Huang Y, Oppewal H. Why consumers hesitate to shop online: An experimental choice analysis of grocery shopping and the role of delivery fees. Int J Retail Distrib Manag. 2006;34(4-5):334–353. doi:10.1108/09590550610660260

41. De Kervenoael R, Soopramanien D, Hallsworth A, Elms J. Personal privacy as a positive experience of shopping: An illustration through the case of online grocery shopping. Int J Retail Distrib Manag. 2007;35(7):583–599. doi:10.1108/09590550710755958

42. Ellis-Chadwick F, Doherty NF, Anastasakis L. E-strategy in the UK retail grocery sector: A resource-based analysis. Manag Serv Qual. 2007;17(6):702–727. doi:10.1108/09604520710835019

43. Gan L, He S, Huang T, Tan J. A comparative analysis of online grocery pricing in Singapore. Electron Commer Res Appl. 2007;6(4):474–483. doi:10.1016/j.elerap.2007.02.006

44. McClatchey J, Cattell K, Michell K. The impact of online retail grocery shopping on retail space: A Cape Town case study. Facilities. 2007;25(3-4):115–126. doi:10.1108/02632770710729700

45. Dias MB, Locher D, Li M, El-Deredy W, Lisboa PJG. The value of personalised recommender systems to e-business: a case study. In: Proceedings of the 2008 ACM Conference on Recommender Systems. RecSys ’08. Association for Computing Machinery; 2008:291–294. doi:10.1145/1454008.1454054

46. Rettie R, Robinson H, Radke A, Ye X. CAQDAS: A supplementary tool for qualitative market research. Qual Mark Res. 2008;11(1):76–88. doi:10.1108/13522750810845568

47. Lim H, Widdows R, Hooker NH. Web content analysis of e-grocery retailers: A longitudinal study. Int J Retail Distrib Manag. 2009;37(10):839–851. doi:10.1108/09590550910988020

48. Milkman KL, Rogers T, Bazerman MH. I’ll have the ice cream soon and the vegetables later: A study of online grocery purchases and order lead time. Mark Lett. 2010;21(1):17–35. doi:10.1007/s11002-009-9087-0

49. Chu J, Arce-Urriza M, Cebollada-Calvo JJ, Chintagunta PK. An Empirical Analysis of Shopping Behavior Across Online and Offline Channels for Grocery Products: The Moderating Effects of Household and Product Characteristics. J Interact Mark. 2010;24(4):251–268. doi:10.1016/j.intmar.2010.07.004

50. Milkman KL, Beshears J. Mental accounting and small windfalls: Evidence from an online grocer. J Econ Behav Organ. 2009;71(2):384–394. doi:10.1016/j.jebo.2009.04.007

51. Wang RJH, Malthouse EC, Krishnamurthi L. On the Go: How Mobile Shopping Affects Customer Purchase Behavior. J Retail. 2015;91(2):217–234. doi:10.1016/j.jretai.2015.01.002

52. İlhan BY, ioğlu TE. Effect of women’s labor market status on online grocery shopping, the case of Turkey. Eurasian Bus Rev. 2015;5(2):371–396. doi:10.1007/s40821-015-0029-x

53. Fisher G, Kotha S. HomeGrocer.com: Anatomy of a failure. Bus Horiz. 2014;57(2):289–300. doi:10.1016/j.bushor.2013.12.002

54. Schnellbaecher C, Behr J, Leonhaeuser IU. Potential of Online Food Shopping Opportunity to Relieve mothers’ Everyday Life Food Routines? Ernahrungs Umsch. 2015;62(11):M626-+.

55. Sathiyaraj. Consumer perception towards online grocery stores, Chennai-Indian Journals. https://www.indianjournals.com/ijor.aspx?target=ijor:zijmr&volume=5&issue=6&article=003

56. Chintagunta PK, Chu J, Cebollada J. Quantifying transaction costs in online/off-line grocery channel choice. Mark Sci. 2012;31(1):96–114. doi:10.1287/mksc.1110.0678

57. Campo K, Breugelmans E. Buying Groceries in Brick and Click Stores: Category Allocation Decisions and the Moderating Effect of Online Buying Experience. J Interact Mark. 2015;31:63–78. doi:10.1016/j.intmar.2015.04.001

58. Breugelmans E, Campo K. Effectiveness of In-Store Displays in a Virtual Store Environment. J Retail. 2011;87(1):75–89. doi:10.1016/j.jretai.2010.09.003

59. Pozzi A. Shopping cost and brand exploration in online grocery. Am Econ J Microecon. 2012;4(3):96–120. doi:10.1257/mic.4.3.96

60. Galante N, López EG, Monroe S. The future of online grocery in Europe. McKinsey Co. Published online 2013:22–31.

61. Dawes J, Nenycz-Thiel M. Comparing retailer purchase patterns and brand metrics for in-store and online grocery purchasing. J Mark Manag. 2014;30(3-4):364–382. doi:10.1080/0267257X.2013.813576

62. de Kervenoael R, Elms J, Hallsworth A. Influencing online grocery innovation: Anti-choice as a trigger for activity fragmentation and multi-tasking. Futures. 2014;62:155–163. doi:10.1016/j.futures.2014.04.004

63. de Kervenoael R, Hallsworth A, Elms J. Household pre-purchase practices and online grocery shopping. J Consum Behav. 2014;13(5):364–372. doi:10.1002/cb.1484

64. Shi SW, Zhang J. Usage experience with decision aids and evolution of online purchase behavior. Mark Sci. 2014;33(6):871–882. doi:10.1287/mksc.2014.0872

65. Melis K, Campo K, Breugelmans E, Lamey L. The Impact of the Multi-channel Retail Mix on Online Store Choice: Does Online Experience Matter? J Retail. 2015;91(2):272–288. doi:10.1016/j.jretai.2014.12.004

66. Muhammad NS, Sujak H, Abd Rahman S. Buying groceries online: the influences of electronic service quality (eServQual) and situational factors. In: Rashid WEW, Muda M, eds. Fifth International Conference on Marketing and Retailing (5th INCOMaR) 2015. Vol 37. ; 2015:379–385. doi:10.1016/S2212-5671(16)30140-X

67. Suel E, Le Vine S, Polak J. Empirical Application of Expenditure Diary Instrument to Quantify Relationships between In-Store and Online Grocery Shopping: Case Study of Greater London. Vol 2496.; 2015:54. doi:10.3141/2496-06

68. Lee D, Jeong H, Cho J, Jeong J, Moon J. Grocery shopping via T-commerce in Korea: New shopping channel adoption behavior based on prior E-commerce experience. Int Food Agribus Manag Rev. 2015;18(2):1–16.

69. MC, Beulens JWJ, Rutters F, et al. The effects of nudges on purchases, food choice, and energy intake or content of purchases in real-life food purchasing environments: a systematic review and evidence synthesis. Nutr J. 2020;19(1):103. doi:10.1186/s12937-020-00623-y

70. Dannenberg P, Fuchs M, Riedler T, Wiedemann C. Digital Transition by COVID-19 Pandemic? The German Food Online Retail. Tijdschr Voor Econ En Soc Geogr J Econ Soc Geogr Rev Geogr Econ Hum Z Okonomische Soz Geogr Rev Geogr Econ Soc. Published online June 19, 2020. doi:10.1111/tesg.12453

71. N, Fraser KT, Arnow C, Mulcahy M, Hille C. Online grocery shopping by NYC public housing residents using supplemental nutrition assistance program (SNAP) benefits: A service ecosystems perspective. Sustain Switz. 2020;12(11). doi:10.3390/su12114694

72. Blitstein JL, Frentz F, Jilcott Pitts SB. A Mixed-method Examination of Reported Benefits of Online Grocery Shopping in the United States and Germany: Is Health a Factor? J Food Prod Mark. 2020;26(3):212–224. doi:10.1080/10454446.2020.1754313

73. Šontaitė-Petkevičienė M. Customer-based brand equity creation for online grocery stores. In: Vol 35. ; 2020:199–206. doi:10.22616/rrd.26.2020.029

74. Van Droogenbroeck E, Van Hove L. Intra-household task allocation in online grocery shopping: Together alone. J Retail Consum Serv. 2020;56. doi:10.1016/j.jretconser.2020.102153

75. Jilcott Pitts SB, Ng SW, Blitstein JL, et al. Perceived Advantages and Disadvantages of Online Grocery Shopping among Special Supplemental Nutrition Program for Women, Infants, and Children (WIC) Participants in Eastern North Carolina. Curr Dev Nutr. 2020;4(5):nzaa076. doi:10.1093/cdn/nzaa076

76. Van Droogenbroeck E, Van Hove L. Adoption of Online Grocery Shopping: Personal or Household Characteristics? J Internet Commer. 2017;16(3):255–286. doi:10.1080/15332861.2017.1317149

77. Ghai S, Tripathi S. Perceived benefits & risks of online grocery shoppingrole of cognitive influences. Indian J Public Health Res Dev. 2019;10(4):29–35. doi:10.5958/0976-5506.2019.00659.4

78. Bauerová R. ONLINE GROCERY SHOPPING IS A PRIVILEGE OF MILLENNIAL CUSTOMERS. STILL TRUTH IN COVID-19 PANDEMIC? Nakupování Potravin Online Jako Výsada Milen Je Stá Pravda V Období Pandemie Covidu-19. 2021;21(1):15–28.

79. Chenarides L, Grebitus C, Lusk JL, Printezis I. Food consumption behavior during the COVID-19 pandemic. Agribus N Y N. Published online December 15, 2020. doi:10.1002/agr.21679

80. Chen X, Kassas B, Gao Z. Impulsive purchasing in grocery shopping: Do the shopping companions matter? J Retail Consum Serv. 2021;60:102495. doi:10.1016/j.jretconser.2021.102495

81. Skotnicka M, Karwowska K, Kłobukowski F, Wasilewska E, Małgorzewicz S. Dietary Habits before and during the COVID-19 Epidemic in Selected European Countries. Nutrients. 2021;13(5). doi:10.3390/nu13051690

82. Ben Hassen T, El Bilali H, Allahyari MS. Impact of covid-19 on food behavior and consumption in qatar. Sustain Switz. 2020;12(17):1–18. doi:10.3390/su12176973

83. Khan M, Ahmed E. Online Grocery Shopping and Consumer Perception: A Case of Karachi Market in Pakistan. Social Science Research Network; 2018. Accessed June 23, 2021. https://papers.ssrn.com/abstract=3671215

84. Zheng Q, Chen J, Zhang R, Wang HH. What factors affect Chinese consumers’ online grocery shopping? Product attributes, e-vendor characteristics and consumer perceptions. China Agric Econ Rev. 2020;12(2):193–213. doi:10.1108/CAER-09-2018-0201

85. Berg J, Henriksson M. In search of the ‘good life’: Understanding online grocery shopping and everyday mobility as social practices. J Transp Geogr. 2020;83. doi:10.1016/j.jtrangeo.2020.102633

86. Lacko A, Ng SW, Popkin B. Urban vs. Rural Socioeconomic Differences in the Nutritional Quality of Household Packaged Food Purchases by Store Type. Int J Environ Res Public Health. 2020;17(20):7637. doi:10.3390/ijerph17207637

87. Zatz LY, Moran AJ, Franckle RL, et al. Comparing Online and In-Store Grocery Purchases. J Nutr Educ Behav. 2021;53(6):471-479. doi:10.1016/j.jneb.2021.03.001

88. Hillen J, Fedoseeva S. E-commerce and the end of price rigidity? J Bus Res. 2021;125:63–73. doi:10.1016/j.jbusres.2020.11.052

89. Aparicio D, Metzman Z, Rigobon R. The Pricing Strategies of Online Grocery Retailers. National Bureau of Economic Research; 2021. doi:10.3386/w28639

90. van Ewijk BJ, Steenkamp JBEM, Gijsbrechts E. The Rise of Online Grocery Shopping in China: Which Brands Will Benefit? J Int Mark. 2020;28(2):20–39. doi:10.1177/1069031X20914265

91. Bhatnagar P, Scarborough P, Kaur A, Dikmen D, Adhikari V, Harrington R. Are food and drink available in online and physical supermarkets the same? A comparison of product availability, price, price promotions and nutritional information. Public Health Nutr. 2021;24(5):819–825. doi:10.1017/S1368980020004346

92. Ayadi K, Muratore I. Digimums’ online grocery shopping: the end of children’s influence? Int J Retail Distrib Manag. 2020;48(4):348–362. doi:10.1108/IJRDM-09-2019-0291

93. Ben Hassen T, El Bilali H, Allahyari MS, Berjan S, Fotina O. Food purchase and eating behavior during the COVID-19 pandemic: A cross-sectional survey of Russian adults. Appetite. 2021;165:105309. doi:10.1016/j.appet.2021.105309

94. Van Droogenbroeck E, Van Hove L. Triggered or evaluated? A qualitative inquiry into the decision to start using e-grocery services. Int Rev Retail Distrib Consum Res. Published online 2019. doi:10.1080/09593969.2019.1655085

95. Olzenak K, French S, Sherwood N, Redden JP, Harnack L. How Online Grocery Stores Support Consumer Nutrition Information Needs. J Nutr Educ Behav. 2020;52(10):952–957. doi:10.1016/j.jneb.2020.07.009

96. Campo K, Lamey L, Breugelmans E, Melis K. Going Online for Groceries: Drivers of Category-Level Share of Wallet Expansion. J Retail. Published online 2020. doi:10.1016/j.jretai.2020.05.003

97. Rogus S, Guthrie JF, Niculescu M, Mancino L. Online Grocery Shopping Knowledge, Attitudes, and Behaviors Among SNAP Participants. J Nutr Educ Behav. 2020;52(5):539–545. doi:10.1016/j.jneb.2019.11.015

98. Martinez O, Tagliaferro B, Rodriguez N, Athens J, Abrams C, Elbel B. EBT Payment for Online Grocery Orders: a Mixed-Methods Study to Understand Its Uptake among SNAP Recipients and the Barriers to and Motivators for Its Use. J Nutr Educ Behav. 2018;50(4):396–402.e1. doi:10.1016/j.jneb.2017.10.003

99. Wang Y, Xu R, Schwartz M, Ghosh D, Chen X. COVID-19 and Retail Grocery Management: Insights from a Broad-Based Consumer Survey. IEEE Eng Manag Rev. 2020;48(3):202–211. doi:10.1109/EMR.2020.3011054

100. Sousa R, Horta C, Ribeiro R, Rabinovich E. How to serve online consumers in rural markets: Evidence-based recommendations. Bus Horiz. 2020;63(3):351–362. doi:10.1016/j.bushor.2020.01.007

101. Kang C, Moon J, Kim T, Choe Y. Why consumers go to online grocery: Comparing vegetables with grains. In: Vol 2016-March. ; 2016:3604–3613. doi:10.1109/HICSS.2016.450

102. Stones C. Online food nutrition labelling in the UK: how consistent are supermarkets in their presentation of nutrition labels online? Public Health Nutr. 2016;19(12):2175–2184. doi:10.1017/S1368980015003110

103. Munson J, Tiropanis T, Lowe M. Online Grocery Shopping: Identifying Change in Consumption Practices. Vol 10673 LNCS.; 2017:211. doi:10.1007/978-3-319-70284-1_16

104. Seitz C, Pokrivčák J, Tóth M, Plevný M. Online grocery retailing in Germany: an explorative analysis. J Bus Econ Manag. 2017;18(6):1243–1263. doi:10.3846/16111699.2017.1410218

105. Wills B, Arundel A. Internet-enabled access to alternative food networks: A comparison of online and offline food shoppers and their differing interpretations of quality. Agric Hum Values. 2017;34(3):701–712. doi:10.1007/s10460-017-9771-2

106. Banerjee T, Banerjee A. Web content analysis of online grocery shopping web sites in India. Int J Bus Anal. 2018;5(4):61–73. doi:10.4018/IJBAN.2018100104

107. Bauerová R. Consumers’ decision-making in online grocery shopping: The impact of services offered and delivery conditions. Acta Univ Agric Silvic Mendel Brun. 2018;66(5):1239–1247. doi:10.11118/actaun201866051239

108. Lazarus MA, Tandon PS, Otten JJ. Examining Relationships between Food Procurement Characteristics and Nutritional Quality in Washington State Child Care Settings. Child Obes Print. 2018;14(6):429–439. doi:10.1089/chi.2018.0090

109. Richards TJ, Rabinovich E. The long-tail of online grocery shopping. Agribusiness. 2018;34(3):509–523. doi:10.1002/agr.21553

110. Loketkrawee P, Bhatiasevi V. Elucidating the Behavior of Consumers toward Online Grocery Shopping: The Role of Shopping Orientation. J Internet Commer. 2018;17(4):418–445. doi:10.1080/15332861.2018.1496390

111. Rishi B, Pradeep H, Vishwanathan M. Hesitation to adoption in the e-grocery retailing in an emerging market. Int J Bus Innov Res. 2018;15(1):99–118. doi:10.1504/IJBIR.2018.088476

112. Weber AN, Badenhorst-Weiss JA. The ‘new’ bricks-and-mortar store: An evaluation of website quality of online grocery retailers in BRICS countries. Afr J Sci Technol Innov Dev. 2018;10(1):85–97. doi:10.1080/20421338.2017.1394957

113. Cebollada J, Chu Y, Jiang Z. Online Category Pricing at a Multichannel Grocery Retailer. J Interact Mark. 2019;46:52–69. doi:10.1016/j.intmar.2018.12.004

114. Hirogaki M. Frequency of retail services, membership fees and real store shopping experience: Analysing consumer preferences. Int J Bus Glob. 2019;23(3):367–382. doi:10.1504/IJBG.2019.102911

115. Kian TP, Loong ACW, Fong SWL. Customer Purchase Intention on Online Grocery Shopping. Int J Acad Res Bus Soc Sci. 2019;8(12):Pages 1579–1595. doi:10.6007/IJARBSS/v8-i12/5260

116. Šarkovská K, Chytková Z. Benefits and pitfalls of online grocery shopping as perceived by the consumers: Evidence from the czech republic. Privred Kretanja Ekon Polit. 2019;27(2):35–58. doi:10.15179/pkiep.27.2.2

117. Singh R. Why do online grocery shoppers switch or stay? An exploratory analysis of consumers’ response to online grocery shopping experience. Int J Retail Distrib Manag. 2019;47(12):1300–1317. doi:10.1108/IJRDM-10-2018-0224

118. Alaimo LS, Fiore M, Galati A. How the COVID-19 pandemic is changing online food shopping human behaviour in Italy. Sustain Switz. 2020;12(22):1–18. doi:10.3390/su12229594

119. Brand C, Schwanen T, Anable J. ‘Online Omnivores’ or ‘Willing but struggling’? Identifying online grocery shopping behavior segments using attitude theory. J Retail Consum Serv. 2020;57. doi:10.1016/j.jretconser.2020.102195

120. de la Llave Montiel MA, López F. Spatial models for online retail churn: Evidence from an online grocery delivery service in Madrid. Pap Reg Sci. 2020;99(6):1643–1665. doi:10.1111/pirs.12552

121. Frank DA, Peschel AO. Sweetening the Deal: The Ingredients that Drive Consumer Adoption of Online Grocery Shopping. J Food Prod Mark. 2020;26(8):535–544. doi:10.1080/10454446.2020.1829523

122. Hao N, Wang HH, Zhou Q. The impact of online grocery shopping on stockpile behavior in Covid-19. China Agric Econ Rev. 2020;12(3):459–470. doi:10.1108/CAER-04-2020-0064

123. Hillen J. Online food prices during the COVID-19 pandemic. Agribus N Y N. Published online December 3, 2020. doi:10.1002/agr.21673

124. Jribi S, Ben Ismail H, Doggui D, Debbabi H. COVID-19 virus outbreak lockdown: What impacts on household food wastage? Environ Dev Sustain. Published online April 19, 2020:1–17. doi:10.1007/s10668-020-00740-y

125. Klepek M, Bauerová R. Why do retail customers hesitate for shopping grocery online? Technol Econ Dev Econ. 2020;26(6):1444–1462. doi:10.3846/tede.2020.13970

126. Oncini F, Bozzini E, Forno F, Magnani N. Towards food platforms? An analysis of online food provisioning services in Italy. Geoforum. 2020;114:172–180. doi:10.1016/j.geoforum.2020.06.004

127. Zafar F, Perepu I. BigBasket’s Struggle with Covid-19. IUP J Supply Chain Manag. 2020;17(3):50–67.

128. Bezirgani A, Lachapelle U. Qualitative Study on Factors Influencing Aging Population’s Online Grocery Shopping and Mode Choice When Grocery Shopping in Person. Transp Res Rec. 2021;2675(1):79–92. doi:10.1177/0361198120964790

129. Droogenbroeck EV, Van Hove L. Adoption and usage of E-grocery shopping: A context-specific UTAUT2 model. Sustain Switz. 2021;13(8). doi:10.3390/su13084144

130. Ellison B, McFadden B, Rickard BJ, Wilson NLW. Examining Food Purchase Behavior and Food Values During the COVID-19 Pandemic. Appl Econ Perspect Policy. 2021;43(1):58–72. doi:10.1002/aepp.13118

131. Ferrante MJ, Goldsmith J, Tauriello S, Epstein LH, Leone LA, Anzman-Frasca S. Food Acquisition and Daily Life for U.S. Families with 4-to 8-Year-Old Children during COVID-19: Findings from a Nationally Representative Survey. Int J Environ Res Public Health. 2021;18(4). doi:10.3390/ijerph18041734

132. Harnack L, Redden J, French S, et al. Designing Online Grocery Stores to Support Healthy Eating for Weight Loss. Public Health Nutr. Published online February 26, 2021:1–33. doi:10.1017/S1368980021000896

133. Khan SA, Ahmad S, Jamshed M. IoT-enabled services in online food retailing. J Public Aff. 2021;21(1). doi:10.1002/pa.2150

134. Rummo PE, Higgins I, Chauvenet C, Vesely A, Jaacks LM, Taillie L. A Standardized Guide to Developing an Online Grocery Store for Testing Nutrition-Related Policies and Interventions in an Online Setting. Int J Environ Res Public Health. 2021;18(9). doi:10.3390/ijerph18094527

135. Zatz LY, Moran AJ, Franckle RL, et al. Comparing shopper characteristics by online grocery ordering use among households in low-income communities in Maine. Public Health Nutr. Published online May 25, 2021:1–6. doi:10.1017/S1368980021002238

136. Zimmer M, McElrone M, Anderson Steeves ET. Feasibility and Acceptability of a “Click & Collect” WIC Online Ordering Pilot. J Acad Nutr Diet. Published online July 1, 2021:S2212-2672(21)00339-7. doi:10.1016/j.jand.2021.05.015

137. Kim S, Lee KI, Heo SY, Noh SC. Identifying Food Deserts and People with Low Food Access, and Disparities in Dietary Habits and Health in Korea. Int J Environ Res Public Health. 2020;17(21). doi:10.3390/ijerph17217936

138. Hillen J. Psychological pricing in online food retail. Br Food J. Published online 2021. doi:10.1108/BFJ-09-2020-0847

139. Gil R, Korkmaz E, Sahin O. Can free-shipping hurt online retailers? Quant Mark Econ. 2020;18(3):305–342. doi:10.1007/s11129-020-09225-8

140. Who Are The Top Online Grocery Shoppers In Europe? Accessed March 23, 2023. https://www.forbes.com/sites/ninaangelovska/2019/04/27/who-are-the-top-online-grocery-shoppers-in-europe/?sh=879d2065b460

141. Top 100 E-commerce in Europe. Accessed March 23, 2023. https://www.retail-index.com/E-commerceretail.aspx

142. Van Droogenbroeck E, Van Hove L. Are the Time-Poor Willing to Pay More for Online Grocery Services? When ‘No’Means ‘Yes.’ J Theor Appl Electron Commer Res. 2022;17(1):253–290.

143. Chester J, Kopp K, Montgomery KC. Does Buying Groceries Online Put SNAP Participants at Risk? How to Protect Health, Privacy, and Equity. Center for Digital Democracy; 2020. Accessed April 25, 2023. https://www.democraticmedia.org/sites/default/files/field/public-files/2020/cdd_snap_report_ff_0.pdf

144. Zhang Q, Patel P, Lowery CM. Protecting Low-Income Consumers in the Era of Digital Grocery Shopping: Implications for WIC Online Ordering. Nutrients. 2023;15(2):390. doi:10.3390/nu15020390

145. Facts and figures 2021 - Internet use in urban and rural areas. Accessed March 23, 2023. https://www.itu.int/itu-d/reports/statistics/2021/11/15/internet-use-in-urban-and-rural-areas

146. Rummo PE, Roberto CA, Thorpe LE, Troxel AB, Elbel B. Age-Specific Differences in Online Grocery Shopping Behaviors and Attitudes among Adults with Low Income in the United States in 2021. Nutrients. 2022;14(20):4427. doi:10.3390/nu14204427

147. Kvalsvik F. Understanding the role of situational factors on online grocery shopping among older adults. J Retail Consum Serv. 2022;68:103009. doi:10.1016/j.jretconser.2022.103009

148. Peck J, Childers TL. If I touch it I have to have it: Individual and environmental influences on impulse purchasing. J Bus Res. 2006;59(6):765–769. doi:10.1016/j.jbusres.2006.01.014

149. Muruganantham G, Bhakat RS. A Review of Impulse Buying Behavior. Int J Mark Stud. 2013;5(3):p149. doi:10.5539/ijms.v5n3p149

150. Caspi CE, Lenk K, Pelletier JE, et al. Association between store food environment and customer purchases in small grocery stores, gas-marts, pharmacies and dollar stores. Int J Behav Nutr Phys Act. 2017;14(1):76. doi:10.1186/s12966-017-0531-x

151. Chrisinger BW, Kallan MJ, Whiteman ED, Hillier A. Where do U.S. households purchase healthy foods? An analysis of food-at-home purchases across different types of retailers in a nationally representative dataset. Prev Med. 2018;112:15–22. doi:10.1016/j.ypmed.2018.03.015

152. Stern D, Robinson WR, Ng SW, Gordon-Larsen P, Popkin BM. US Household Food Shopping Patterns: Dynamic Shifts Since 2000 And Socioeconomic Predictors. Health Aff (Millwood). 2015;34(11):1840–1848. doi:10.1377/hlthaff.2015.0449

153. Mah CL, Luongo G, Hasdell R, Taylor NGA, Lo BK. A Systematic Review of the Effect of Retail Food Environment Interventions on Diet and Health with a Focus on the Enabling Role of Public Policies. Curr Nutr Rep. 2019;8(4):411–428. doi:10.1007/s13668-019-00295-z

154. Fan L, Ellison B, Wilson NLW. What Food waste solutions do people support? J Clean Prod. 2022;330:129907. doi:10.1016/j.jclepro.2021.129907

155. US EPA O. Preventing Wasted Food At Home. Published April 18, 2013. Accessed May 9, 2023. https://www.epa.gov/recycle/preventing-wasted-food-home

156. Krebs-Smith SM, Guenther PM, Subar AF, Kirkpatrick SI, Dodd KW. Americans Do Not Meet Federal Dietary Recommendations. J Nutr. 2010;140(10):1832–1838. doi:10.3945/jn.110.124826

157. Juul F, Parekh N, Martinez-Steele E, Monteiro CA, Chang VW. Ultra-processed food consumption among US adults from 2001 to 2018. Am J Clin Nutr. 2022;115(1):211–221. doi:10.1093/ajcn/nqab305

158. Martínez Steele E, Popkin BM, Swinburn B, Monteiro CA. The share of ultra-processed foods and the overall nutritional quality of diets in the US: evidence from a nationally representative cross-sectional study. Popul Health Metr. 2017;15(1):6. doi:10.1186/s12963-017-0119-3

159. Mokdad AH, Ballestros K, Echko M, et al. The State of US Health, 1990-2016: Burden of Diseases, Injuries, and Risk Factors Among US States. JAMA. 2018;319(14):1444–1472. doi:10.1001/jama.2018.0158

160. Jardim TV, Mozaffarian D, Abrahams-Gessel S, et al. Cardiometabolic disease costs associated with suboptimal diet in the United States: A cost analysis based on a microsimulation model. PLOS Med. 2019;16(12):e1002981. doi:10.1371/journal.pmed.1002981

161. Etminani-Ghasrodashti R, Hamidi S. Online shopping as a substitute or complement to in-store shopping trips in Iran? Cities. 2020;103:102768. doi:10.1016/j.cities.2020.102768

162. Avery J, Steenburgh TJ, Deighton J, Caravella M. Adding Bricks to Clicks: On the Role of Physical Stores in a World of Online Shopping. NIM Mark Intell Rev. 2013;5(2):28–33. doi:10.2478/gfkmir-2014-0015

163. Monteiro CA, Cannon GJ. The role of the transnational ultra-processed food industry in the pandemic of obesity and its associated diseases: problems and solutions. World Nutr. 2019;10(1):89–99. doi:10.26596/wn.201910189-99

164. Rummo PE, Roberto CA, Thorpe LE, Troxel AB, Elbel B. Effect of Financial Incentives and Default Options on Food Choices of Adults With Low Income in Online Retail Settings: A Randomized Clinical Trial. JAMA Netw Open. 2023;6(3):e232371. doi:10.1001/jamanetworkopen.

